# Cerebrovascular and cardiovascular autonomic regulation in sickle cell patients with white matter lesions

**DOI:** 10.1101/2023.04.18.23288776

**Authors:** Christophe Ferreira De Matos, Pierre Cougoul, Oana Maria Zaharie, Marc Kermorgant, Anne Pavy-Le Traon, Celine Gales, Jean-Michel Senard, Mathilde Strumia, Fabrice Bonneville, Nathalie Nasr

**Author notes:** Corresponding author : Christophe Ferreira De Matos.

## Abstract

**Background:** The prevalence of asymptomatic white matter lesions (WML) in patients with sickle cell disease (SCD) has been described to be very frequent in young adults. Cerebrovascular regulation and cardiovascular autonomic regulation, more specifically the sympatho-vagal balance can be altered in SCD.

In this study we assessed the association between WML, cerebrovascular regulation and sympatho-vagal balance in SCD.

**Method:** Adults with no history of stroke from a cohort of SCD patients were prospectively evaluated for, cerebrovascular regulation using Mx for autoregulation, breath holding test for cerebrovascular reactivity and cerebral arterial compliance calculated from arterial blood pressure and cerebral velocities. Sympatho-vagal balance was assessed using heart rate variability parameters. WML was assessed with MRI using Fazekas score grading and the presence of lacunar lesions.

**Results:** Forty-one patients (F/M:25/16) were included. Median age was 37.5 (range 19-65). Twenty-nine (70,7%) patients had SS genotype, 7 patients (17,1%) had SC genotype and 5 (12,2%) patients had Sß° genotype. Among the 41 patients included, 11 patients had WML (26,8%). Patients with WML were significantly older (44.5 vs 30.6 years; p<0.001), had a lower HF (HF 157 ms^2^ vs HF 467.6 ms^2^; p<0.005) and impaired cerebral arterial compliance (CaBVR 15.4 vs 37.3 cm^3^/mmHg; p<0.014). Cerebral blood flow velocities, reactivity to breath holding test and cerebral autoregulation parameters did not significantly differ between the two groups.

**Conclusions:** Lower parasympathetic activity and impaired cerebral arterial compliance were associated with WML in adults with SCD. This could potentially yield to a better understanding of pathophysiological parameters leading to premature cerebrovascular ageing in SCD patients.

## Introduction

Sickle cell disease (SCD) is a common genetic hemoglobinopathy, resulting in chronic hemolysis, inflammation and endothelial dysfunction. Neurovascular damage remains the leading cause of stroke in children and is a major cause of morbidity and mortality in the sickle cell patients (1). White matter lesions (WML) and silent lacunar are frequent in SCD. Their prevalence has been described to be as high as 53% by in HbSS or HbSß° patients with a median age of 30 (2). They are defined by white matter hyperintensities (WMH) on magnetic resonance imaging (MRI). Both stroke and silent cerebral infarction are associated with a high risk of recurrence, cognitive impairment and impaired social and professional quality of life (3,4). The pathophysiological mechanisms are complex and partially understood. Chronic anemia induces tissular hypoxia leading to a secondary increase in cerebral blood flow to maintain adequate oxygenation (5). Cerebral autoregulation is expected to compensate through cerebrovascular dilation in a context associated with potentially defective vasomotor reserve due to low NO availability (6,7). Autonomic nervous system (ANS) is a major component of cerebrovascular homeostasis. Indeed, a defective ANS is an independent predictor of both cardiovascular and neurovascular events in patients without SCD (8– 10). Impaired parasympathetic function has been described in SCD patients (11–15).

The aim of the study was to evaluate the association between alterations in ANS, cerebrovascular regulation and the prevalence of WML in adult patients with SCD.

## Methods

### Study population

Data were collected prospectively from adult sickle cell patient cohort at the University Hospital Center of Toulouse in France, between January 2017 and April 2021. SCD was defined by SS, SC or composite heterozygous genotype (Sß° thalassemia). Exclusion criteria were age <18, the absence of transcranial doppler (TCD) acoustic window and a history of stroke.

Chronic treatment was defined by the use of Hydroxyurea, transfusion or both. Laboratory tests were performed during routine outpatient visits. Patients had assessment of BRS, HRV, cerebral hemodynamics, and 3T MRI as part the follow-up.

All patients provided written informed consent. The research protocol was approved by the institutional ethics committee (registration number 09-815). STROBE checklist items were adequately addressed.

### BRS and HRV measurement

BRS was measured at rest in time domain using the x-correlation method (16) with patients lying supine in a quiet, temperature-controlled room. Inter-beat intervals were derived from the time in milliseconds between sequential R-peaks on a 3-lead ECG monitor and ABP from continuous monitoring using Finapres (Finometer, Cardionet, France). BRS sensitivity was calculated in ms/mmHg.

HRV was measured in frequency domain using fast Fourier transform (9). Measurements were performed in low frequency (LF:0.04– 0.15 Hz) and high frequency (HF:0.15–0.40 Hz). The LF power reflected sympathetic and parasympathetic function whereas HF power reflected solely parasympathetic function. The ratio LF/HF indicated sympathovagal balance.

### Assessment of cerebral hemodynamics

All patients had transcranial color-coded ultrasound (TCCS) to assess velocities in proximal intracranial arteries including the MCA, before they had continuous monitoring of MCA velocities TCD during 10 to 20 minutes. A 2MHz probe was fixed with a headframe to the temporal acoustic window unilaterally at a depth of 50-55mm (ATYS Looki Rhône-Alpes, France). Analog outpouts from TCD and Finapres were synchronized and exported to ICM+ software for the assessment of cerebral autoregulation using Mx (19) -higher Mx indicating more impaired autoregulation-, arterial cerebral compliance was assessed using the method described by Czosnyka et al. (21) and the assessment of cerebrovascular reactivity to CO2 using a 30 seconds breath-holding test.

#### Breath holding test

Voluntary apnea was performed during 30 seconds. ABP was continuously measured to detect Valsalva maneuver. ETCO2 was monitored to detect micro-inspirations. This test was performed 3 times and the best performed test in terms of absence of micro-inspirations and Valsava maneuver was used for analysis. The results in terms of increase of velocities were given as percentage (17).

### Cerebral MRI

Each patient underwent cerebral MRI (Philips Achieva 3.0T MRI), with quantification of WML assessed by the Fazekas score and/or the presence of at least one lacunar infarct (18). Each MRI was independently reviewed by 2 radiologists blinded the cerebral hemodynamics assessment. This evaluation allowed to define two groups : one without any WML, the other with WML graded with Fazekas score ≥ 1 and/or the presence of lacunar infarcts.

### Statistical analysis

Distribution of continuous variables were presented as mean and standard deviation; categorical variables were described by numbers and proportions. Student T test (or Wilcoxon test if the conditions were not met) was used for continuous variables and the Chi^2^ test (or Fisher’s exact test if the conditions were not met) for categorical variables. Statistical analysis was conducted with R software (version 4.0.3).

## Results

### General characteristics

Forty-one patients were included. Median age was 37.5 years (range 19-65). There were 25 (61.0%) women and 16 (39.0%) men. Twenty-nine (70.7%) patients had SS genotype, 7 patients (17.1%) had SC genotype and 5 (12.2%) patients had Sß° genotype. Amongst the 41 patients, 11 had (26.8%) WML. Patients with WML were significantly older (44.5 vs 30.6 years; p<0.001). There was no difference in sex ratio, BMI, SCD genotype, arterial blood pressure, hemoglobin level, hemoglobin F percentage, hemolysis parameters and treatment between the two groups (Table 1).

**Table 1.** General characteristics of SCD adult patients with and without WML on MRI.

| Characteristics | Absence of WML<br>n = 30 | Presence of WML<br>n = 11 | p-value |
| --- | --- | --- | --- |
| Age - yr | 30.6 (± 10.1) | 44.5 (± 10.4) | < **0.001** |
| Sex - n |  |  | 0.383 |
| Female (%) | 20 (66.7%) | 5 (45.5%) |  |
| Male (%) | 10 (33.3%) | 6 (54.5%) |  |
| Genotype |  |  | 0.643 |
| SS (%) | 21 (70.0%) | 8 (72.7%) | 1.000 |
| SC (%) | 6 (20.0%) | 1 (9.1%) | 0.651 |
| Sb° (%) | 3 (10.0%) | 2 (18.2%) | 0.598 |
| Hemoglobin (g/dL) | 9.1 (± 1.9) | 9.0 (± 1.9) | 0.780 |
| Reticulocytes (G/L) | 226.6 (± 106.4) | 223.3 (± 78.1) | 0.914 |
| Platelets (G/L) | 291.5 (± 100.0) | 373.5 (± 141.8) | 0.100 |
| Bilirubin (mmol/l) | 35.6 (± 20.3) | 32.1 (± 17.3) | 0.746 |
| LDH (UI/l) | 397.1 (± 165.4) | 472.4 (± 259.4) | 0.471 |
| Hb F (%) | 6.6 (± 6.3) | 9.1 (± 7.9) | 0.281 |
| Smoke |  |  | 1.000 |
| Yes/no | 4 (13.3%) / 26 (86.7%) | 1 (9.1%) / 10 (90.9%) |  |
| Cannabis |  |  | 0.299 |
| Yes/no | 5 (16.7%) / 25 (83.3%) | 0 (0.0%) / 11 (100%) |  |
| BMI | 22.2 (± 3.3) | 22.9 (± 5.1) | 0.883 |
| Systolic BP (mmHg) | 117.0 (± 11.5) | 124.5 (± 12.8) | 0.109 |
| Diastolic BP (mmHg) | 65.0 (± 9.9) | 67.4 (± 12.8) | 0.542 |
| HR (bpm) | 82.7 (± 12.2) | 78.1 (± 13.0) | 0.323 |
| Chronic treatment |  |  |  |
| Yes/no | 18 (60.0%) / 12 (40.0%) | 5 (45.4%) / 6 (54.5%) | 0.498 |
| Hydroxyurea | 15 (50.0%) | 3 (27.3%) | 0.345 |

LDH: Lactates deshydrogenase, HbF: Hemoglobin F percentage, BMI: Body Mass Index, BP: Blood Pressure, HR: Heart Rate.

### Autonomic nervous system

Patients with WML had a lower HF (HF 157.1 ms^2^ vs HF 467.6 ms^2^; p<0.005). There were no difference between the two groups for BRS (Table 2).

**Table 2.**
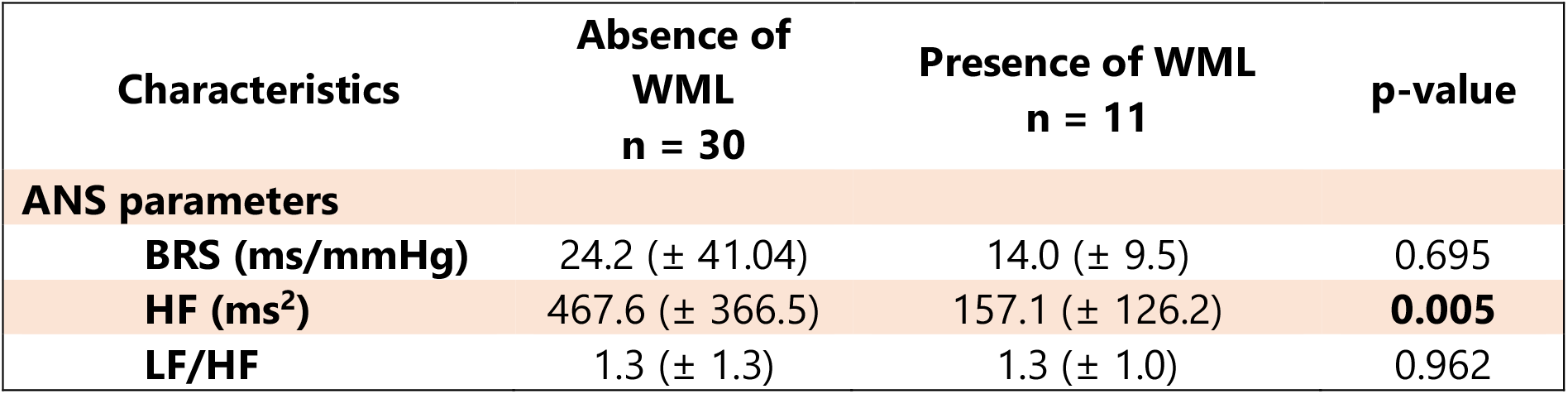
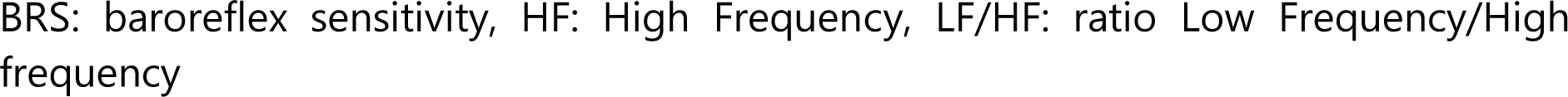
Autonomic nervous system parameters of SCD patients with and without WML.

| Characteristics | Absence of WML<br>n = 30 | Presence of WML<br>n = 11 | p-value |
| --- | --- | --- | --- |
| ANS parameters |  |  |  |
| BRS (ms/mmHg) | 24.2 (± 41.04) | 14.0 (± 9.5) | 0.695 |
| HF (ms<sup>2</sup>) | 467.6 (± 366.5) | 157.1 (± 126.2) | **0.005** |
| LF/HF | 1.3 (± 1.3) | 1.3 (± 1.0) | 0.962 |

BRS: baroreflex sensitivity, HF: High Frequency, LF/HF: ratio Low Frequency/High frequency

### Cerebrovascular regulation parameters

Cerebral blood flow velocities of middle cerebral artery did not significantly differ between the two groups (105.3 vs 115.5 cm/sec; p=0.381), nor did BHT and Mx (Table 3).

**Table 3.** Cerebovascular regulation parameters of patients with SCD with and without WML.

| Characteristics | Absence of WML<br>n = 30 | Presence of WML<br>n = 11 | p-value |
| --- | --- | --- | --- |
| **Cerebral autoregulation parameters** |  |  |  |
| **CBFV (cm/s)** | 105.3 (± 25.4) | 115.5 (± 34.2) | 0.381 |
| **BHT (%)** | 43.4 (± 11.7) | 46.3 (± 13.0) | 0.582 |
| **Mx** | 0.2 (± 0.2) | 0.2 (± 0.4) | 0.949 |
| **CaBVR**<br>**(cm<sup>3</sup>/mmHg)** | 37.3 (± 22.8) | 15.4 (± 19.1) | **0.014** |

CBFV: Systolic Cerebral Blood Flow Velocity of middle cerebral artery, BHT: Breath holding Test, Correlation coefficient Mx, CaBVR: Cerebral Arterial compliance.

Patient with WML had lower cerebral arterial compliance compared with patients without WML (CaBVR 15.4 vs 37.3 cm^3^/mmHg; p<0.014)

### Association between WML and the autonomic nervous system parameters

BRS did not significantly differ between the two groups (14,0 ms/mmHg in the group with WML vs. 24,2 ms/mmHg in the group without WML; p=0.695).

For HRV parameters, univariate logistic regression showed an inverse relationship between the presence of WML and/or lacunar infarcts and HF (for each increase of 100 units in HF, OR: 0.46; CI 0.21-1.00; p=0.050). In multivariate analysis, after adjustment for age there was no significant statistical association with WML (for each increase of 100 units in HF, OR: 0,59; CI 0,29-1,19; p=0,085)

The area under the receiver operating characteristic curve for the model with HF as a single predictor was estimated at 0.857. For the model with age as a single predictor, area under the receiver operating characteristic curve was estimated at 0.876. For the age and HF model, area under the receiver operating characteristic curve was 0.946. For the age, HF and CaBVR model, area under the receiver operating characteristic curve was 1.(Figure 1).

**Figure 1.**
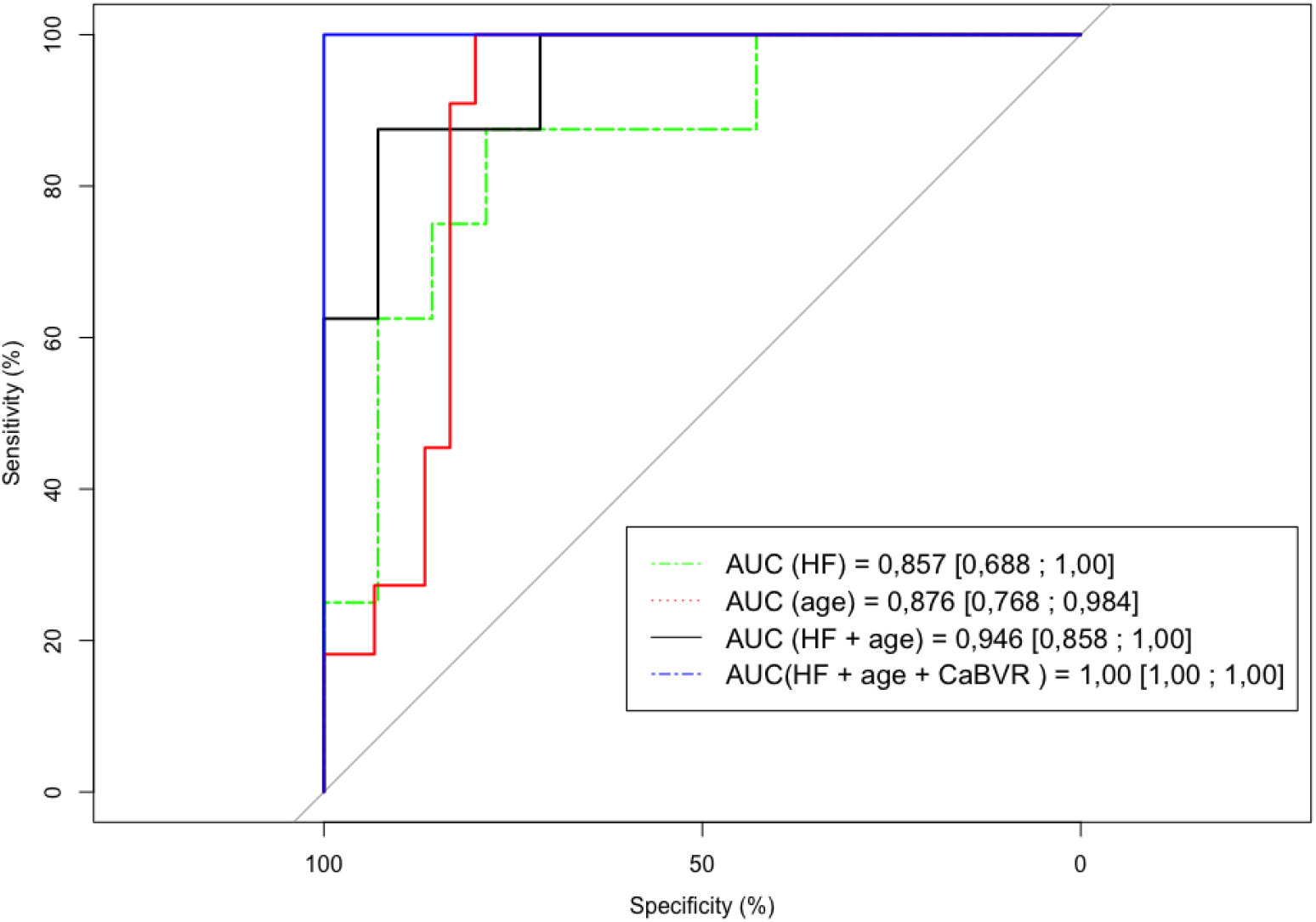
Receiver operating characteristic (ROC) curves for the regression models predicting the WML in SCD patients from : HF alone, Age alone, HF + Age and Age + HF + CaBVR.

## Discussion

In our study the presence of WML in patients with SCD was associated with altered parasympathetic function and lower cerebral arterial compliance. WML corresponds to white matter changes (WMC) in SCD patients and are observed in cerebrovascular ageing in the general population. To our knowledge, this is the first study assessing cerebrovascular regulation and cardiovascular autonomic regulation in relation with WML in patients with SCD.

In our study, WML was defined by the presence of white matter hyperintensity (WMH) or/and lacunar infarcts. In a meta-analysis, Debette et al. showed that WMH is one of the earliest markers of small vessel disease and is associated with increased risk of dementia, stroke and death in patients without SCD (19). The prevalence of WML is more likely to be present in older patients. It affected 5% of people aged 50 to almost 100% of people older than 90 years in the general population (20). In our study, WML affected 26,8% of patients with a mean age of 44 years old.

Previous studies showed contradictory results concerning the risk factors for WML microangiopathy in SCD patients. The study by Debaun et al. reported that male sex, high systolic blood pressure, low level of hemoglobin were associated with a high risk of WML (21). Calvet et al, showed that low fetal hemoglobin percentage was associated with white matter changes, but not low level of hemoglobin (22).

Impairment of parasympathetic function has been reported in SCD, but its association with WML has not been described yet (11,23). Imbalance of the autonomic nervous system, particularly activation of the sympathetic system has been associated with platelet aggregation, inflammation, arrhythmia and cardiac ischemia in patients without SCD (24). This imbalance of the ANS with higher sympathetic activity and lower parasympathetic activity is one of the many pathophysiological mechanisms hypothesized in SCD. However, the exact mechanisms at place remain unknown (13). Tracey et al. reported that a decrease in parasympathetic activity might promote inflammation, by inhibiting acetylcholine-mediated cytokine release from leukocytes (25). Some authors suggested that the predominance of sympathetic activity could be a trigger for promoting the transition from the basal state to vaso-occlusive crisis mediated by vasoconstriction (23,26).

Cerebral arterial compliance is the property of an artery to expand and contract passively with changes in arterial blood pressure. However, due to limited access to the cerebral arteries, cerebral arterial compliance has not been assessed so far in studies on SCD. Some studies have evaluated cerebral arterial compliance by cerebral MRI, defined as the ratio of intracranial volume change to cerebrospinal fluid pressure gradient during cardiac cycle (27,28). However, this provides a snapshot of compliance and not a continuous assessment of this variable in time-domain. Carrera et al. developed a new mathematical model, based on continuous monitoring of arterial blood pressure using photo-plethysmograph and cerebral blood flow velocities using TCD to assess cerebral arterial compliance (29,30). Arterial compliance has been shown to be altered on the side of stenosis/occlusion and its impairment correlated with the severity of the internal carotid artery disease (30). The assessment of cerebral arterial compliance has not been described in SCD patients. In our study, we showed that WML was associated with an altered cerebral arterial compliance. This new method of assessing cerebral arterial compliance is more physiological, sensitive, and probably shows earlier changes than the alteration of the breath holding test. In our study, the cerebral arterial compliance was reduced whilst the breath holding test did not significantly differ between the two groups. These results might be explained by the potent capacity of CO2 retention from BHT to elicit dilation of the distal cerebrovascular even when little remaining spontaneous vasodilation capacity remains.

The pathophysiological mechanisms of WML, appear to be multifactorial. Chronic ischemia, lower NO bioavailability, inflammation (chronic hemolysis and impairment of parasympathetic system), vascular remodeling due to endotheliopathy and cardiovascular stress lead to WML (31).

This study has limitations. First, a relatively limited number of patients were included and secondly there was no control group. This cohort was however characterized by a systemic follow up in patients with no patent cerebrovascular disease. Cerebral MRI as well as cerebral hemodynamics assessment was performed once as part of an annual check-up. Also, MRI were blindly reviewed by two independent neuroradiologists blinded to the results of the assessment of cerebral hemodynamics.

This study provides new insight into the association between cardiovascular and cerebrovascular hemodynamic characteristics and the prevalence of WML in adult patients with SCD. Firstly, lower HF indicating lower parasympathetic activity was associated with the presence of WML. Secondly, altered cerebral arterial compliance in sickle cell patients was associated with WML unlike the breath holding test result, and from that point of view seemed to be a more sensitive parameter.

Our results are monocentric and need to be confirmed in larger multicentric studies. They could potentially yield a better understanding of pathophysiological parameters that lead to premature cerebrovascular ageing in patients with SCD.

Several preventive cardiovascular therapies, including non-pharmacological treatment including physical activity, can improve cerebrovascular function and parasympathetic activity. HF and CaBVR monitoring can help monitor the impact of therapies that aim to slow down cerebrovascular ageing in SCD patients.

## Data Availability

All data referred to in the manuscript were obtained with the consentement of patients

